# Analytical validation of HepatoPredict kit to assess hepatocellular carcinoma prognosis prior to a liver transplantation

**DOI:** 10.1101/2023.05.30.23290711

**Authors:** Maria Gonçalves-Reis, Daniela Proença, Laura P. Frazão, João L. Neto, Sílvia Silva, Hugo Pinto-Marques, José B. Pereira-Lea, Joana Cardoso

**Affiliations:** - Ophiomics – Precision Medicine, Lisbon, Portugal; - Hepato-Biliary-Pancreatic and Transplantation Centre, Curry Cabral Hospital, Centro Hospitalar Universitário de Lisboa Central, Lisbon, Portugal; – Chronic Diseases Research Center (CEDOC), NOVA Medical School, Universidade NOVA de Lisboa (NMS/UNL), Lisbon,Portugal

**Keywords:** HepatoPredict, Liver cancer, Hepatocellular carcinoma, Liver transplantation, Analytical validation, Multi-target genomic assay, Prognostic test

## Abstract

**Background:** The best curative treatment for hepatocellular carcinoma (HCC) is liver transplant (LT), but the limited number of organs available for LT dictates strict eligibility criteria. Despite this patient selection stringency, current criteria often fail in pinpointing patients at risk of HCC relapse and in identifying good prognosis patients that could benefit from a LT. HepatoPredict kit was developed and clinically validated to forecast the benefit of LT in patients diagnosed with HCC. By combining clinical variables and a gene expression signature in an ensemble of machine learning algorithms, HepatoPredict stratifies HCC patients according to their risk of relapse after LT.

**Methods:** Aiming at the characterization of the analytical performance of HepatoPredict kit in terms of sensitivity, specificity and robustness, several variables were tested which included reproducibility between operators and between RNA extractions and RT-qPCR runs, interference of input RNA levels or varying reagent levels. The described methodologies, included in the HepatoPredict kit, were tested according to analytical validation criteria of multi-target genomic assays described in guidelines such as ISO201395-2019, MIQE, CLSI-MM16, CLSI-MM17, and CLSI-EP17-A. Furthermore, a new retrained version of the HepatoPredict algorithms is also presented and tested.

**Results:** The results of the analytical performance demonstrated that the HepatoPredict kit performed within the required levels of robustness (*p* > 0.05), analytical specificity (inclusivity ≥ 95 %), and sensitivity (LoB, LoD, linear range, and amplification efficiency between 90 – 110 %). The introduced operator, equipment, input RNA and reagents into the assay had no significant impact on HepatoPredict classifier results. As demonstrated in a previous clinical validation, a new retrained version of the HepatoPredict algorithm still outperformed current clinical criteria, in the accurate identification of HCC patients that more likely will benefit from a LT.

**Conclusions:** Despite the variations in the molecular and clinical variables, the prognostic information obtained with HepatoPredict kit and does not change and can accurately identify HCC patients more likely to benefit from a LT. HepatoPredict performance robustness also validates its easy integration into standard diagnostic laboratories.

## Background

Primary liver cancer is the 6^th^ most diagnosed cancer and the third leading cause of cancer death worldwide [1]. Hepatocellular carcinoma (HCC) comprises 75-85 % of primary liver cancer cases and it is associated with chronic infection with hepatitis B virus (HBV) or hepatitis C virus (HCV), aflatoxin-contaminated foods, heavy alcohol intake, excess body weight, type 2 diabetes, and smoking [1]. About 30 % of HCC cases are considered for treatment with curative intent [2] which involves liver transplantation (LT) or surgical resection [2,3]. In contrast with surgical resection, LT treats the HCC as well as the underlying cirrhosis, reducing the patient’s risk of death within the first 2 years of diagnosis [3]. However, due to the shortage of liver donors, several different criteria mainly based on tumor burden and protein biomarkers such as alpha-fetoprotein (AFP) and des-γ carboxyprothrombin (DCP), have been developed for the identification of HCC patients most likely to benefit from LT [4–13]. Nevertheless, the limitations of these criteria are currently under discussion, mainly because they exclude patients with an underlying good prognosis who can benefit from a LT and include bad prognosis patients that will not benefit from the surgery [14,15].

The HepatoPredict kit intends to predict which patients diagnosed with HCC have a good prognosis and thus will benefit from a LT. This is achieved by combining three clinical variables (tumor number, size of the largest nodule, and total tumor volume) and a gene expression signature (includes *DPT, CLU, CAPNS1,* and *SPRY2 genes*) and a proprietary algorithm. In short, the HepatoPredict kit can extract RNA from formalin-fixed paraffin-embedded (FFPE) HCC samples and to perform gene expression analysis through real-time quantitative reverse transcription polymerase chain reaction (RT-qPCR) technology. The RT-qPCR results are then combined with the clinical data using a machine learning algorithm that returns three different values concerning the predictive value (Class I and II) or its absence (Class 0) [16] (Figure 1).

**Figure 1.**
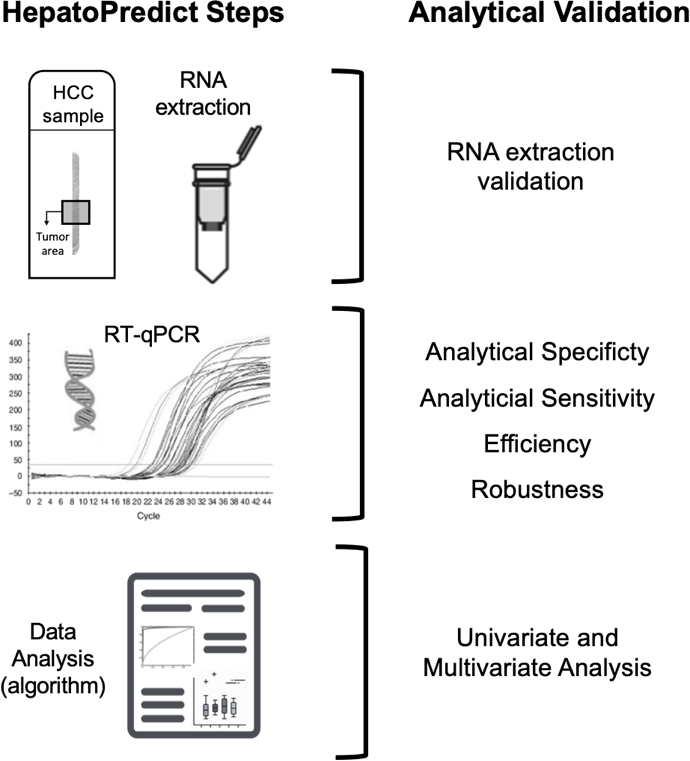
– HepatoPredict kit Analytical Validation. The HepatoPredict kit intends to predict tumor recurrence after a liver transplant in patients with HCC. The HepatoPredict kit uses FFPE HCC samples from which the RNA is extracted and subsequently used as a template in 1-step RT-qPCR reactions targeting reference genes (*RPL13A*, *GAPDH*, *TBP*), genes of interest (*DPT, CLU, CAPNS1, SPRY2*) and a genomic DNA control (*Chr3*). Gene expression levels of the genes of interest are normalized to the geometric mean of the reference genes and combined with clinical variables through an algorithm. The analytical validation of the HepatoPredict kit comprised several different assays performed at different steps of the kit: validation of the RNA extraction methodology, analytical specificity, sensitivity, efficiency, and robustness of RT-qPCR reactions and univariate and multivariate analysis of the HepatoPredict algorithm.

In a previous study [16], using a retrospective clinical validation cohort of patients diagnosed with HCC, we have demonstrated that HepatoPredict outperforms the Milan [4], University of California San Francisco (UCSF) [6], Up-to-seven [11], alpha fetoprotein (AFP) [7], Metroticket 2.0 [5], Total Tumour Volume (TTV) [13], and TTV AFP [12] criteria in the selection of patients suitable for LT ^16^. Apart from its clinical utility, the technical performance of a prognostic test used in the diagnostic setting must also be verified through its capacity to generate specific, sensitive, and robust data under standard laboratory conditions (analytical validation). The aim of this work is to present evidence of the analytical validation of the HepatoPredict kit based on comprehensive technical studies (Figure 1). This validation also included the re-training of the HepatoPredict algorithm using a retrospective cohort of 162 patients diagnosed with HCC and submitted to LT.

## Methods

### Samples

In this study, HCC samples preserved as FFPE tissue were used. FFPE HCC samples were acquired from four different suppliers: Biobank IRBLleida (PT20/00021), integrated in the Spanish National Biobanks Network and Xarxa de Bancs de Tumors de Catalunya (XBTC) sponsored by Pla Director d’Oncología de Catalunya; Biobank ISABIAL, integrated in the Spanish National Biobanks Network and in the Valencian Biobanking Network; and biorepositories from Amsbio (US) and Biotech (US). All samples were processed following standard operating procedures with the appropriate approval of the Ethical and Scientific Committees. Moreover, clinical samples from a retrospective clinical study approved by the ethics authorities and taking place in the Curry Cabral Hospital (Lisbon, Portugal), were also used. All HCC FFPE samples were acquired either sectioned with 3-5 μm thickness or as paraffin blocks that were then cut in 3-5 μm thick slices using a microtome (Leica SM2010R Sliding Microtome, Leica Biosystems) and mounted on a glass slide.

### Histopathologic analysis

Prior to RNA extraction, HCC FFPE samples were analyzed by a certified pathologist using an hematoxylin and eosin (H&E) stained tumor section. FFPE HCC slides (3 μm thick) were first deparaffinized and stained using Harris Hematoxylin solution (#3801561E, Leica Biosystems, Richmond, USA. The slide was then counterstained with Eosin Y solution (#2801601, Leica Biosystems, Richmond, USA). Finally, slides were dehydrated in increasing alcohol concentrations, cleared in xylene (#28973, VWR, Alfragide, Portugal), and mounted using a xylene-based mounting medium (#107961, Merck, Darmstadt, Germany). After H&E staining, slides were observed under an optical upright microscope (Panther L, #1100104600142, Motic^Ò^).

### RNA extraction

For RNA extraction, an HCC area mimicking a needle biopsy was delimited in two sequential 5 μm slides. Samples were initially deparaffinized and the RNA was extracted using the RNeasy FFPE Kit (#73504, Qiagen, Hilden, Germany), according to the manufacturer’s instructions with two exceptions: proteinase K cell lysis and final elution volume.

### DNA extraction

DNA was extracted from HCC FFPE samples using the QIAamp DNA FFPE Tissue Kit (#56404, Qiagen, Hilden, Germany) in accordance with the manufacturer’s instructions.

### RNA extraction method validation

For the validation of the RNA extraction method, 87 FFPE HCC samples (from the four different suppliers) were used. Each sample was tested in duplicate and by two different operators. Moreover, mirror sections of each sample were used to reduce the variability between operators. Immediately after extraction, RNA was stored at −20 °C until further usage or used straight away in RT-qPCR reactions (conditions described below) targeting *RPL13A* (reference gene) and *Chr3* (genomic DNA control) to analyze the integrity [17] and the purity of the RNA samples, respectively.

### RT-qPCR reactions

1-step RT-qPCR reactions were performed as previously described [16]. The QuantStudio Design & Analysis Software v1.5.1 software was used for data acquisition and analysis. For gene expression normalization, the geometric mean of the cycle threshold (Cq) of the reference genes (*RPL13A, GAPDH,* and *TBP*) was subtracted from the Cq values of the genes of interest (*DPT, CLU, CAPNS1,* and *SPRY2*).

### Primer Specificity

RT-qPCR products were sequenced via Sanger sequencing outsourced to Eurofins (https://eurofinsgenomics.eu/en/custom-dna-sequencing/gatc-services/supremerun-tube/). In total, 16 different solutions (forward and reverse for 8 targets) were sent to Eurofins. Regarding RT-qPCR products, a 2-step RT-qPCR reaction was performed using the SuperScript™ VILO™ cDNA Synthesis Kit (#11754050, Thermo Fisher Scientific, Bleiswijk, Netherlands) to synthetize cDNA and the Invitrogen^TM^ Platinum^TM^ SuperFi^TM^ PCR Master Mix with the SuperFi^TM^ GC Enhancer (#12358010, Thermo Fisher Scientific, Bleiswijk, Netherlands) in qPCR. An RNA pool (composed of 8 different FFPE HCC samples) was used as template. The size of each RT-qPCR product was assessed by electrophoresis in a 4 % agarose gel (#G401004, Thermo Fisher Scientific, Bleiswijk, Netherlands), using a DNA ladder (#10488096, Thermo Fisher Scientific, Vilnius, Lithuania) and nuclease-free water (#129114, Qiagen, Hilden, Germany) as a negative control in an electrophoresis system (#G8300, Thermo Fisher Scientific, Vilnius, Lithuania).

### RT-qPCR inclusivity

Different FFPE HCC samples were used as templates in RT-qPCR reactions targeting all the genes included in HepatoPredict kit. Each RT-qPCR reaction was performed in duplicate and by two different operators. For each sample, each operator used the same batch of extracted RNA or DNA. Both nucleic acid extraction and RT-qPCR reactions were performed as described above.

### Limit of Detection (LoD) determination

For LoD determination, a pool composed of 8 FFPE HCC samples was used. The samples composing the pool reflected high and low expression levels of each target and were associated with a bad prognosis (recurrence, n = 4) and a good prognosis (no recurrence, n = 4). Both pools, of DNA and RNA, were created using the same samples. DNA pool was directly used for serial dilutions (at least 11 per target) and the RNA pool was diluted 1:4 to create the starting sample for the serial dilutions. In total, 21 replicates were done for each dilution (triplicates in each of the 7 RT-qPCR reactions), per lot number of reagents, on three different days (2-3 RT-qPCR reactions per day). For each target, all reactions were performed by the same operator with the same equipment. RT-qPCR reactions using reagents from different lots were analyzed separately. Data was analyzed in accordance with the Probit model, which implied the creation of a regression representing the probability vs log_2_ dilution for each target assuring at least 3 dilutions with hit rates within 0.10 - 0.90 and at least one exceeding 0.95. Moreover, to minimize the influence of the model limit ranges of probability, dilutions with a 100 % fail or success rate were included in each analysis. LoDs were independently calculated for each lot and the maximum LoD (concentration) was taken as the reported value for the measurement procedure. To determine the Cq value associated with the LoD, a linear regression was applied between the Cq values and the log_2_(dilution factor). All the log_2_(dilution factor) until the one immediately after the LoD were considered. The values outside the confidence interval (CI) at 99 % were considered outliers and were removed. Linear, quadratic, and cubic polynomial functions were fitted to the Cq values using log_2_ dilution values. If none of the non-linear coefficients was different from zero, the target was considered linear (*GAPDH*, *TBP* and *Chr3*). Otherwise, the absolute difference between the model that best fits the data (smallest mean squared error) and the linear model was calculated. When the difference was less than 1 Cq value, the target was considered linear (*RPL13A, DPT, CAPNS1, CLU* and *SPRY2*).

### Linearity

The linear range of each target included in the HepatoPredict kit was determined for RT-qPCR reactions using FFPE HCC samples (previously used for LoD determination) and reference materials (#636690, Takara, Saint Germain en Laye, France) to cover a broad range of nucleic acids concentrations in linearity determination. For that, seven serial dilutions of reference RNA were used with 3 replicates per dilution and at least eleven dilutions of nucleic acids pools, obtained from HCC FFPE samples, were used with 7 replicates per dilution. Finally, the Cq values and the dilution factors were plotted in a base 2 logarithmic graph and R^2^ (> 0.90) was calculated for all targets.

### Amplification efficiency

The reaction efficiency was calculated for each target included in the HepatoPredict kit. It was determined from the slope of the log-linear portion of each target curve: amplification efficiency = (2^-1/slope^ - 1) x 100.

### Robustness of RT-qPCR reactions

Plackett and Burman tables [18] were used to design the robustness assay: alterations in the concentrations (± 30 %) of the master mix (#A15300, Thermo Fisher Scientific, Bleiswijk, Germany), primers and probes were implemented as well as different final reaction volumes (± 5 %) and annealing temperatures (± 1 °C), as demonstrated in Table 1. Two independent assays per each target were performed using the same sample pool (see LoD) in triplicate and all reactions were performed using sample concentration near the LoD (RNA pool serial dilution 2^-2^ and DNA pool without further dilutions). Three standard conditions were incorporated in the assay for data analysis: standard (STD) (no changes), STD1 (-1 °C annealing temperature), and STD2 (+1 °C annealing temperature).

**Table 1.** – Design of the robustness assay for the HepatoPredict kit.

| Factor | Conditions |  |  |  |  |  |  |  |  |  |  |
| --- | --- | --- | --- | --- | --- | --- | --- | --- | --- | --- | --- |
|  | STD | STD1 | STD2 | 1 | 2 | 3 | 4 | 5 | 6 | 7 | 8 |
| PCR Equipment | A | A | A | A | A | A | A | A | A | A | A |
| [Master Mix] | NC | NC | NC | + 30 % | + 30 % | - 30 % | - 30 % | + 30 % | + 30 % | - 30 % | - 30 % |
| [Primers] | NC | NC | NC | NC | - 30 % | NC | + 30 % | NC | - 30 % | NC | + 30 % |
| [Probe] | NC | NC | NC | NC | - 30 % | + 30 % | NC | - 30 % | NC | NC | + 30 % |
| Final Reaction Volume | NC | NC | NC | - 5 % | - 5 % | + 5 % | + 5 % | + 5 % | + 5 % | - 5 % | - 5 % |
| Annealing Temperature | NC | - 1 °C | + 1 °C | + 1 °C | - 1 °C | + 1 °C | - 1 °C | - 1 °C | + 1 °C | - 1 °C | + 1 °C |

STD – standard. NC – no changes regarding the standard protocol.

### Determination of Cq values below LoD and within the linear range for each target

Serial dilutions of the reference RNA (#636690, Takara, Saint Germain en Laye, France) were used as templates for RT-qPCR reactions targeting all genes included in the HepatoPredict kit. The Cq values above the LoD and outside the linear range for each target were identified and the maximum acceptable Cq value for each target was determined.

### Precision studies

The conditions under which repeated measurements were made determine the type of precision being analyzed – reproducibility (daily, lot-to-lot, operator, and inter-assay) and repeatability. For the daily reproducibility, for the same sample, assays were performed by the same operator, using the same sample and kit’s lot on 4 different days. Regarding lot-to-lot reproducibility, the same sample was analyzed by the same operator using kits from three different lots. Finally, operator reproducibility was studied by using the same sample with HepatoPredict kits from the same lot but performed by three different operators. For each condition, two HepatoPredict kits were used (two independent assays). Repeatability was measured considering the triplicates of each HepatoPredict kit run. In total, 3 different HCC FFPE samples were studied, thus, 48 HepatoPredict kits were used (16 kits/sample). To further assess inter-assay reproducibility, 15 additional samples were tested in duplicate by different operators, using different lots of the HepatoPredict kit, and on different days (total n = 18).

### HepatoPredict algorithm training

A dataset with 162 patients diagnosed with HCC and submitted to liver transplant was used (*Supplementary File 1*). Concerning the dataset, different models were tested, such as Naive Bayes, support-vector machine (SVM) with different kernel functions, and Extreme Gradient Booster (XGBoost). Moreover, synthetic minority oversampling technique (SMOTE) was also used for data imbalances. Python 3.8 was used with scikit-learn 1.0.2 (1https://scikit-learn.org/stable/), XGBoost 1.6.1 (https://xgboost.readthedocs.io/en/stable), imbalance-learn (https://imbalanced-learn.org/stable/), and Optuna 2.10.0 (https://optuna.org/). Each model was fed with 4 molecular (*DPT, CLU, CAPNS1*, and *SPRY2* gene expression) and 3 clinical variables (tumor number, largest tumor size and total tumor volume). As previously described [16], the algorithm was developed as a two-level predictor.

### HepatoPredict algorithm univariate analysis

The univariate analysis of the HepatoPredict algorithm consisted in calculating the error (i.e., counting each time the algorithm would fail the correct prognosis classification) when altering the Cq mean values of each gene (prior normalization) and varying the normalized Cq values of *DPT, CLU, SPRY2* and *CAPNS1* and the clinical variables (tumor number, diameter of the largest tumor, and total tumor volume). Thus, Cq mean values of the genes of interest (*DPT, CLU, SPRY2* and *CAPNS1*) were replaced by 40 Cq (the maximum number of cycles allowed) and their respective LoD and the Cq means of the reference genes (*RPL13A, GAPDH*, and *TBP*) were removed and replaced by their respective LoD. Regarding the variations of the normalized Cq values (for *DPT, CLU, SPRY2* and *CAPNS1* genes), variations of 0.1 Cq were performed. Alterations in clinical variables included the variations in the tumor number (1-2 units), and in the diameter of the largest tumor (cm) and the total tumor volume (cm^3^) by 2 %.

### HepatoPredict algorithm multivariate analysis

The multivariate analysis of the HepatoPredict algorithm consisted in calculating the error associated with the alteration of more than one variable at a time. Thus, a range for each variable variation was defined (based on algorithm univariate analysis) and random combinations of 2, 3, 4, 5, 6 and 7 altered variables were tested. All possible combinations of variables were performed and for each combination the assay was repeated 10,000 times with random variable alterations within the defined range. Finally, two types of errors were calculated: error type A (between Class I and Class II) and error type B (between Class I or II and Class 0).

### Statistical analysis

Statistical analysis was performed using the R language for Statistical Computing (v 4.1.1) and GraphPad Prism 7 (GraphPad Software, Inc. 2016). For RNA extraction validation, as the data followed a normal distribution, the Paired Student t-test was applied. For robustness assay, Dunn’s multiple comparisons test was applied. Regarding precision, due to data size, a non-parametric test (Friedman test) was used. A *p* < 0.05 was considered statistically significant.

## Results

### Validation of RNA extraction method

For the RNA extraction from HCC FFPE tissues, the RNeasy FFPE kit was used. This RNA extracted method was validated by using 87 HCC FFPE samples handled by two different operators. After extraction, RNA was used as a template in RT-qPCR reactions targeting the *RPL13A* gene and a DNA-specific target (*Chr3*), assuring both the integrity and the purity of the extracted RNA. Each sample was analyzed in duplicate by each operator. Regarding *RPL13A* expression, no significant differences were observed between operators for each sample (*p* = 0.27, Figure 2A) and the mean standard deviation (SD) between samples tested by the two operators was 0.47 Cq (Figure 2D). Moreover, no statistically significant differences were observed between each operator’s duplicates (*p* = 0.99 for operator 1 and *p* = 0.13 for operator 2, Figure 2B and 2C respectively). The mean SD between duplicates of each sample for operator 1 was 0.12 Cq while it was 0.18 Cq for operator 2 (Figure 2D). Regarding *Chr3*, residual genomic DNA (gDNA) contamination (Cq mean > 34) was identified in 8 samples (9.19 %) handled by operator 2 and in one sample (1.15 %) handled by operator 1 (data not shown).

**Figure 2.**
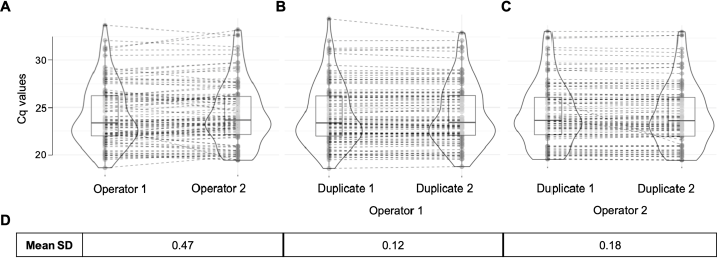
– RNA extraction validation using *RPL13A* gene expression. The RNA extraction method was validated by two different operators (*p* = 0.27, Paired Student’s t-test) **(A)**. Moreover, each operator performed the assays in duplicate: operator 1 (*p* = 0.99, Paired Student’s t-test) **(B)** and operator 2 (*p* = 0.13, Paired Student’s t-test) **(C)**. The mean SD between operators and within operators is also represented **(D)**. For each violin plot (**A**, **B**, and **C**), dots represent Cq mean values **(A)** and Cq values (**B** and **C**) of *RPL13A* gene. Dashed lines represent the correspondence of samples between groups.

### Analytical specificity – Primer specificity and RT-qPCR inclusivity

To demonstrate primers’ uniqueness for each target, primers and RT-qPCR products were sequenced and the specificity of each primer pair was confirmed by aligning the Sanger sequencing electropherograms from each primer with the respective PCR amplicon, as suggested by ISO 20395:2019 and MIQE guidelines [19,20]. Before sequencing, the amplicon size was verified by electrophoresis in an agarose gel (Figure 3A, *RPL13A* depicted as an example). For the *RPL13A* gene, the electrophoresis band corresponded to 75 bp (Figure 3A), in accordance with the expected amplicon size [16]. All the amplified amplicons (for the remaining HepatoPredict targets) corresponded to the expected size [16] and no extra bands of unspecific PCR products were observed (data not shown) confirming the specificity of the primer pairs for the desired target. The sequences of all primers and probes were successfully aligned for all targets included in the HepatoPredict (data not shown) in the DNA sequence displayed in the Sanger electropherogram of the respective PCR amplicon (Figure 3B-D, example for *RPL13A*). Due to the very small size of the amplicons (between 71 and 108 bp [16]) and to limit the baseline noise always present at the beginning and end of Sanger electropherograms, the Sanger sequencing was performed for both forward and reverse strands (Figure 3C and 3D, respectively). This allowed for the successful sequencing of forward and reverse primers and respective probe positions in all HepatoPredict amplicons.

**Figure 3.**
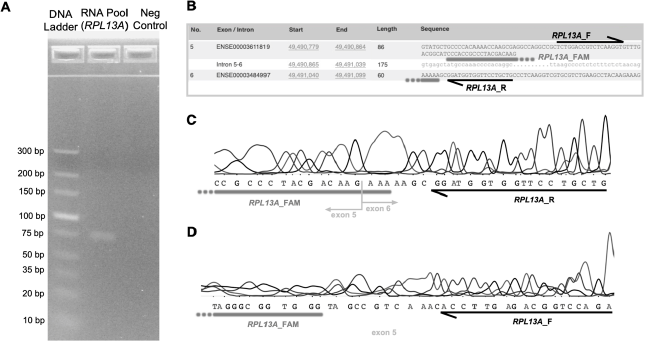
– Primer Specificity (for *RPL13A* as an illustrative example) After a RT-qPCR reaction targeting the *RPL13A* gene, the size of the amplicon was assessed by electrophoresis in an agarose gel (∼75 bp), using a DNA ladder and a negative control (nuclease-free water) **(A)**. ENSEMBL canonical transcript sequence (ENST00000391857.9, RefSeq NM_012423), represented from 5’ to 3’ **(B)**. Alignment of the *RPL13A* forward (*RPL13A*_F) and reverse (*RPL13A*_R) primers and probe (RPL13A_FAM) with the Sanger sequencing electropherogram results for the *RPL13A* amplicon in forward (5’ to 3’) **(C)** and reverse (3’ to 5’) **(D)** directions. The presented image is cropped. The full size original image can be found in *Supplementary File 3*.

The inclusivity of the RT-qPCR reactions included in the HepatoPredict kit was demonstrated as described in CLSI-MM17 guideline [21]. An inclusivity of 100 % was demonstrated for all targets, except for *DPT* which had an inclusivity of 95 % (*Supplementary File 2*).

### Limit of Detection (LoD), Limit of Blank (LoB), Linearity and Efficiency

For LoD, LoB, linearity and efficiency determination of each RT-qPCR reaction included in HepatoPredict kit, an RNA pool of FFPE HCC samples was used in accordance with MM16-A guideline [22]. The LoD, for each of the 8 targets included in the HepatoPredict kit, was determined based on ISO 20395:2019, CLSI-MM17, MIQE, and CLSI-EP17-A guidelines [19–21,23]. The Probit model was used, and Figure 4 exemplifies the application of the model to the *RPL13A* target. The Probit model was applied to two different data sets obtained using different reagent lots (Figure 4A and 4B). The LoD was defined as the lowest concentration of target that could be detected in ≥ 95 % of the samples, as represented in Figure 4A and 4B. To determine the Cq value corresponding to the LoD, a linear regression was performed (Figure 4C). For all targets, Pearson’s correlation coefficient (R^2^) was higher than 0.90 (data not shown), except for *Chr3* (R^2^ = 0.73, data not shown). The highest LoD (nucleic acid concentration) between lots was taken as the reported value for the measurement. The LoD of the 7 RNA targets included in the HepatoPredict kit ranged from 34.75 Cq to 36.89 Cq (Table 2). Regarding *Chr3* the LoD was defined at 33.95 Cq (Table 2).

**Figure 4.**
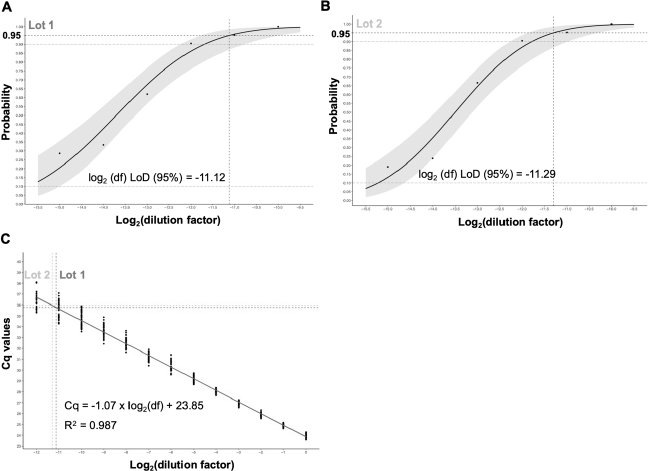
– Estimation of the Limit of Detection (LoD) in Cq values (for *RPL13A* as an illustrative example) The Probit approach was used to determine the LoD of *RPL13A* gene for two different lots of reagents. The LoD was defined as the concentration (log_2_ dilution) at a probability of 95 %. The grey areas represent the confidence interval at 99 % (**A** and **B**). The linear dynamic range was also estimated and the Cq value associated with the LoD was determined **(C)**. For the example of the *RPL13A* gene, the highest LoD was obtained with Lot 1 and the correspondent Cq value was 35.75.

**Table 2.**
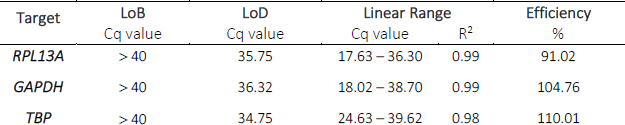
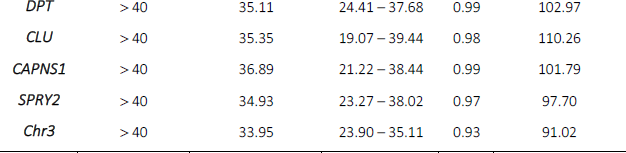
- LoB, LoD, linear range and PCR efficiency of the 8 targets included in HepatoPredict kit.

To apply the Probit model, it is necessary to assume that all blank or negative samples are reported as negative. These assumptions are true for the HepatoPredict kit, in fact, if a valid Cq value was obtained (< 40 cycles) for just one of the replicates of the NTC (no template control) the entire assay was considered invalid. Thus, the LoB was assumed to be zero, i.e., Cq results for the NTC samples, for all valid RT-qPCR reactions, did not cross the threshold within the 40 cycles and were considered “undetermined” (> 40 Cq).

The linear range of each target was also determined in accordance with ISO 20395:2019 [19] with R^2^ = 0.99 for *RPL13A, GAPDH, DPT,* and *CAPNS1*, R^2^ = 0.98 for *TBP* and *CLU*, R^2^ = 0.97 for *SPRY2* and R^2^ = 0.93 for *Chr3*. All targets were linear at least within 24.63 and 35.11 Cq (Table 2). Moreover, the amplification efficiencies, determined in accordance with ISO 20395:2019 and MIQE guidelines [19,20], ranged from 91.02 to 110.26 Cq for all targets (Table 2).

Considering that the HepatoPredict kit analyzes 7 different genes and a DNA-specific target, it is important to assure that all targets can be detected within their linear range and below their LoDs. As represented in Table 3, the maximum RNA input to assure an ideal performance of the HepatoPredict kit is 0.031 ng/μL, corresponding to Cq values of 28.26 for *RPL13A*, 28.34 for *GAPDH*, 33.95 for *TBP*, 34.44 for *DPT*, 29.42 for *CLU*, 31.61 for *CAPNS1*, and 33.96 for *SPRY2*.

**Table 3.** – Acceptable maximum Cq values for each HepatoPredict kit’s target to be detected below their LoD and within the linear range.

| Reference | Targets (Cq mean) |  |  |  |  |  |  |
| --- | --- | --- | --- | --- | --- | --- | --- |
| RNA (ng/μL) | *RPL13A* | *GAPDH* | *TBP* | *DPT* | *CLU* | *CAPNS1* | *SPRY2* |
| 8 | 20.19 | 20.40 | 25.67 | 25.95 | 21.38 | 23.43 | 25.15 |
| 4 | 21.14 | 21.36 | 26.59 | 26.94 | 22.44 | 24.45 | 26.30 |
| 2 | 22.28 | 22.37 | 27.66 | 27.85 | 23.42 | 25.46 | 27.31 |
| 1 | 23.33 | 23.27 | 28.82 | 28.89 | 24.44 | 26.45 | 28.26 |
| 0,5 | 24.06 | 24.28 | 29.76 | 30.02 | 25.40 | 27.68 | 29.26 |
| 0.25 | 25.09 | 25.33 | 30.67 | 31.12 | 26.58 | 28.70 | 30.50 |
| 0.125 | 26.12 | 26.43 | 31.93 | 32.04 | 27.40 | 29.46 | 31.34 |
| 0.063 | 27.06 | 27.47 | 32.76 | 32.47 | 28.60 | 30.75 | 32.58 |
| 0.031 | 28.26 | 28.34 | 33.95 | 34.44 | 29.42 | 31.61 | 33.96 |
| 0.016 | 29.12 | 29.38 | **35.47** | **35.64** | 30.53 | 32.86 | 34.33 |
| 0.008 | 30.31 | 30.34 | **35.31** | **36.68** | 31.79 | 33.89 | **37.39** |
| 0.004 | 31.20 | 31.54 | **36.40** | N/A | 32.67 | 35.02 | **36.80** |
| 0.002 | 32.36 | 32.41 | N/A | **38.06** | 33.68 | **37.12** | N/A |
| 0.00100 | 33.22 | 33.33 | **37.71** | N/A | 34.57 | **37.10** | **38.47** |
| 0.00050 | 34.41 | 34.18 | N/A | N/A | 35.20 | **38.19** | N/A |
| 0.00024 | 35.65 | 36.31 | N/A | **37.19** | **37.15** | **39.17** | N/A |
| 0.00012 | **36.85** | **37.37** | N/A | N/A | N/A | **38.75** | N/A |

Cq mean values represented in bold are above the LoD of each target. Between double lines are represented the acceptable maximum Cq values for all targets assuring that all of them are detected below their LoD and within their linear range: [RNA] = 0.031 ng/μL. N/A – not applicable, Cq > 40.

### Robustness of RT-qPCR reactions

To study the robustness of the RT-qPCR reactions included in the HepatoPredict kit, alterations in the concentrations and volumes of RT-qPCR reagents were performed as suggested in ISO 20395:2019 [19] and represented in Table 1. The Cq mean values of two independent assays obtained for each target under each condition are represented in Figure 5A-C. Conditions with the same annealing temperature were compared with the respective standard condition – conditions 2, 4, 5, and 7 were compared with STD1 (Figure 5B), while conditions 1, 3, 6, and 8 were compared with STD2 (Figure 5C) – and no statistically significant differences were observed (*p* > 0.05). Furthermore, all conditions (from 1 to 8) were compared with the STD condition representing no changes regarding the initial protocol (Figure 5A) and no statistically significant differences were observed (*p* > 0.05).

**Figure 5.**
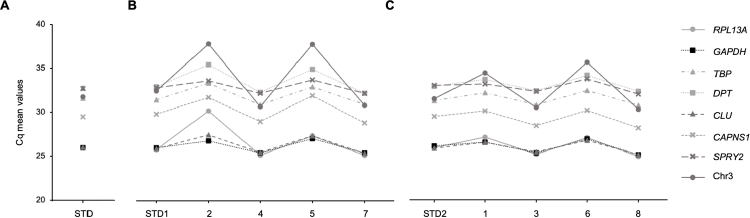
– Robustness of the RT-qPCR reactions included in the HepatoPredict kit. Representation of Cq mean values for each condition (Table 1) of each target included in the HepatoPredict kit. The original condition (STD) **(A)** was compared with all other conditions (1 to 8) and no statistically significant differences were observed (Dunn’s multiple comparisons test). Moreover, conditions with the same annealing temperature were compared with the respective STD condition: **(B)** STD1 was compared with condition 2, 4, 5 and 7 while **(C)** STD2 was compared with conditions 1, 3, 6, and 8. No statistically significant differences were observed (Dunńs multiple comparisons test).

### Precision of the HepatoPredict kit

The precision of the HepatoPredict kit was determined as described in ISO 20395:2019, MIQE, and CLSI- MM17 guidelines [19–21]. Precision data was transduced numerically using imprecision values such as standard deviation (SD) and respective confidence interval (CI) at 95 % (Table 4). The HepatoPredict kit reproducibility was verified by normalizing the gene expression level of the genes of interest (*DPT, CLU, CAPNS2,* and *SPRY2*) to the geometric mean of the reference genes (*RPL13A, GAPDH,* and *TBP*) (Table 4), as described for the standard use of the kit. In general, the SD for daily, lot-to-lot, and operator reproducibility were higher for the *DPT* gene (0.38 – 1.36) when compared with the other genes of interest included in the HepatoPredict kit (0.03 – 0.44). The same was verified for the inter-assay reproducibility (SD calculated between all the independent assays for the same sample) and total SD (square root of the daily, lot-to-lot, and operator variances) (Table 4). Furthermore, while all targets of sample A were associated with higher SD in lot-to-lot reproducibility, sample C presented higher SD values in daily reproducibility. Nevertheless, none of these were observed in sample B, suggesting that the observed variability between independent assays is not dependent on a single factor. In fact, when all varying factors were considered (inter-assay reproducibility and total SD), SD values were similar between both samples. Additionally, inter-assay reproducibility was also determined for 18 different HCC FFPE samples (calculation of the mean SD and respective confidence interval), corroborating the previous results (Table 4). The repeatability was verified for each target included in the HepatoPredict kit – reference genes (*Supplementary File 4*) and genes of interest (Table 4) – and similar SD were obtained for all targets ranging from 0.05 to 0.14 Cq.

**Table 4.**
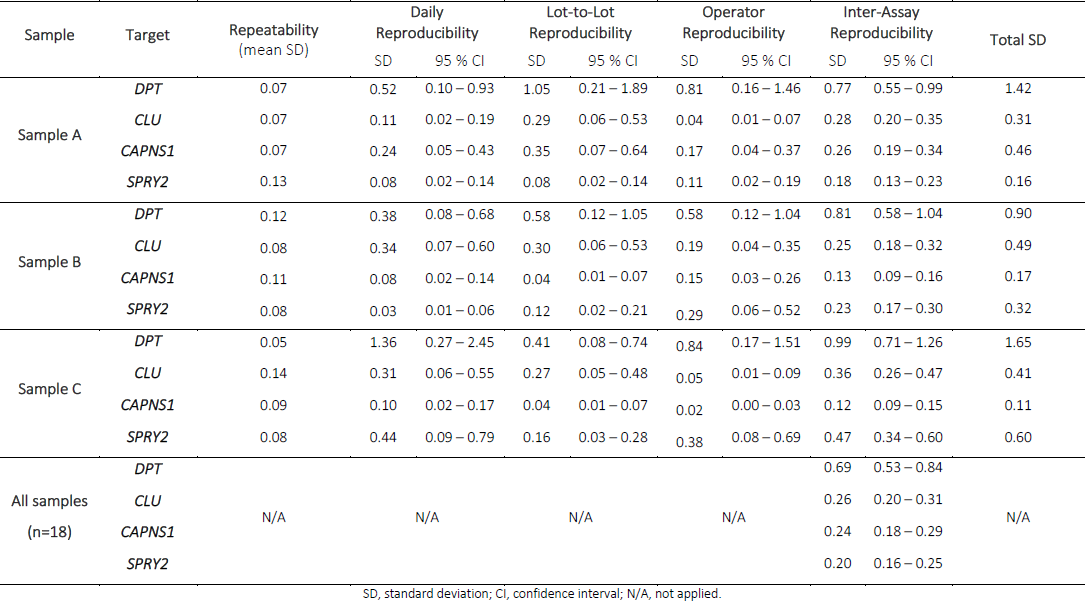
– Reproducibility of the HepatoPredict kit.

| Sample | Target | Repeatability<br>(mean SD) | Daily<br>Reproducibility |  | Lot-to-Lot<br>Reproducibility |  | Operator<br>Reproducibility |  | Inter-Assay<br>Reproducibility |  | Total SD |
| --- | --- | --- | --- | --- | --- | --- | --- | --- | --- | --- | --- |
|  |  |  | SD | 95 % CI | SD | 95 % CI | SD | 95 % CI | SD | 95 % CI |  |
| Sample A | *DPT* | 0.07 | 0.52 | 0.10 – 0.93 | 1.05 | 0.21 – 1.89 | 0.81 | 0.16 – 1.46 | 0.77 | 0.55 – 0.99 | 1.42 |
|  | *CLU* | 0.07 | 0.11 | 0.02 – 0.19 | 0.29 | 0.06 – 0.53 | 0.04 | 0.01 – 0.07 | 0.28 | 0.20 – 0.35 | 0.31 |
|  | *CAPNS1* | 0.07 | 0.24 | 0.05 – 0.43 | 0.35 | 0.07 – 0.64 | 0.17 | 0.04 – 0.37 | 0.26 | 0.19 – 0.34 | 0.46 |
|  | *SPRY2* | 0.13 | 0.08 | 0.02 – 0.14 | 0.08 | 0.02 – 0.14 | 0.11 | 0.02 – 0.19 | 0.18 | 0.13 – 0.23 | 0.16 |
| Sample B | *DPT* | 0.12 | 0.38 | 0.08 – 0.68 | 0.58 | 0.12 – 1.05 | 0.58 | 0.12 – 1.04 | 0.81 | 0.58 – 1.04 | 0.90 |
|  | *CLU* | 0.08 | 0.34 | 0.07 – 0.60 | 0.30 | 0.06 – 0.53 | 0.19 | 0.04 – 0.35 | 0.25 | 0.18 – 0.32 | 0.49 |
|  | *CAPNS1* | 0.11 | 0.08 | 0.02 – 0.14 | 0.04 | 0.01 – 0.07 | 0.15 | 0.03 – 0.26 | 0.13 | 0.09 – 0.16 | 0.17 |
|  | *SPRY2* | 0.08 | 0.03 | 0.01 – 0.06 | 0.12 | 0.02 – 0.21 | 0.29 | 0.06 – 0.52 | 0.23 | 0.17 – 0.30 | 0.32 |
| Sample C | *DPT* | 0.05 | 1.36 | 0.27 – 2.45 | 0.41 | 0.08 – 0.74 | 0.84 | 0.17 – 1.51 | 0.99 | 0.71 – 1.26 | 1.65 |
|  | *CLU* | 0.14 | 0.31 | 0.06 – 0.55 | 0.27 | 0.05 – 0.48 | 0.05 | 0.01 – 0.09 | 0.36 | 0.26 – 0.47 | 0.41 |
|  | *CAPNS1* | 0.09 | 0.10 | 0.02 – 0.17 | 0.04 | 0.01 – 0.07 | 0.02 | 0.00 – 0.03 | 0.12 | 0.09 – 0.15 | 0.11 |
|  | *SPRY2* | 0.08 | 0.44 | 0.09 – 0.79 | 0.16 | 0.03 – 0.28 | 0.38 | 0.08 – 0.69 | 0.47 | 0.34 – 0.60 | 0.60 |
| All samples<br>(n=18) | *DPT* | N/A | N/A | N/A | N/A | N/A | N/A | N/A | 0.69 | 0.53 – 0.84 | N/A |
|  | *CLU* |  |  |  |  |  |  |  | 0.26 | 0.20 – 0.31 |  |
|  | *CAPNS1* |  |  |  |  |  |  |  | 0.24 | 0.18 – 0.29 |  |
|  | *SPRY2* |  |  |  |  |  |  |  | 0.20 | 0.16 – 0.25 |  |

SD, standard deviation; CI, confidence interval; N/A, not applied.

### Univariate and multivariate analysis of the new version of the HepatoPredict algorithm

The HepatoPredict algorithm (V2.0) is a two-level predictor, but the first level (Class I), presents the highest precision and uses the XGboost model (instead of SVM in V1.0) increasing the positive predictive value (PPV) to 96.43 %. The second level (Class II) is a linear SVM model. The variables’ weights within each model are represented in *Supplementary File 5*, with *DPT* and *CLU* gene expression levels and total tumor volume being the most important variables of the HepatoPredict algorithm. Furthermore, the new HepatoPredict algorithm was also compared with other clinical criteria for the identification of HCC patients suitable for liver transplantation (*Supplementary File 6*).

The analytical validation of the new HepatoPredict algorithm consisted in calculating the error (i.e., how many times the correct classification is missed) associated with the alteration of one (univariate) or more (multivariate) variables. Figure 6A represents the error associated with the alteration of the Cq mean values before normalization. The errors were all superior to 10 % demonstrating that to maintain a good HepatoPredict performance, no reference gene can be removed from the assay and the Cq means of the genes cannot be replaced by 40 Cq or its respective LoD. Moreover, the variations allowed for each variable, assuring a maximum error of 5 % (or a maximum variation of 2 Cq or 50 % from the initial value), are represented in Figure 6B-D. In general, it was verified that the *DPT* normalized gene expression level and the total tumor volume (in cm^3^) were the variables that least allowed for alterations (for an error = 5 %, variation of ± 0.4 Cq and ± 12 %, respectively). On the other hand, *CLU* normalized gene expression level and the tumor size (the diameter of the largest tumor in cm) were the variables that tolerated greater variations (± 3 Cq and ± 50 % respectively, while maintaining an error < 5 %) (Figure 6C-D). For these variables, the error at 5 % was not used as a threshold for the multivariable analysis, but instead an acceptable absolute variation value was used (± 2 Cq for *CLU* expression level and ± 50 % for tumor size). Furthermore, while maintaining an error < 5 %, the tumor number was possible to vary in 2 units (Figure 6B) and the normalized gene expression levels of *CAPNS1* and *SPRY2* varied ± 1.7 Cq and ± 1.5 Cq, respectively (Figure 6D).

**Figure 6.**
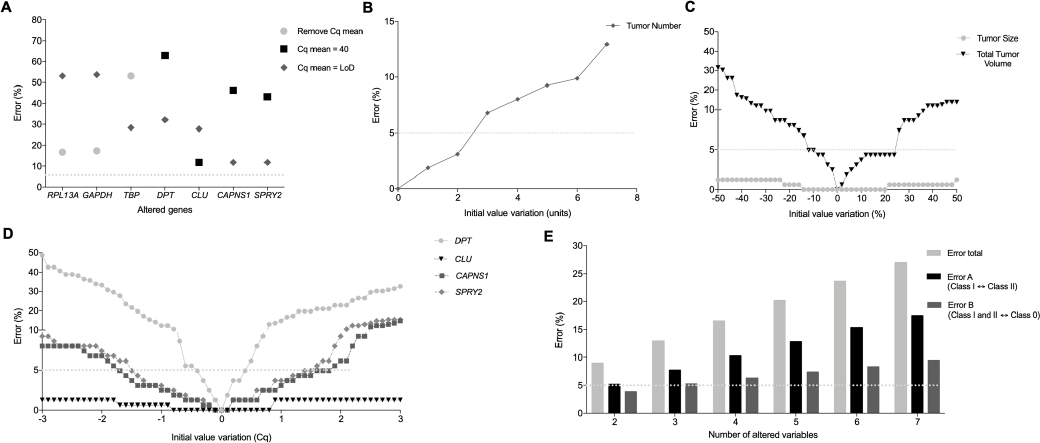
– Robustness of the HepatoPredict algorithm. Representation of errors associated with the univariate alteration of Cq mean values from each molecular variables before normalization **(A)**, with the univariate alteration of the tumor number **(B)**, or tumor size or total tumor volume **(C)**. Errors associated with the univariate alteration of the molecular variables after normalization **(D)**. Multivariate analyses of the HepatoPredict algorithm for distinct combinations of variables, ranging from 2 to 7 variables **(E)**. The dashed line in light grey represents the errors at 5 %.

The HepatoPredict algorithm multivariate analysis was based on the results presented in Figure 6B-D and demonstrated that the error increases with the number of varying variables (Figure 6E). Moreover, two types of errors were analyzed concerning the final HepatoPredict class. Error A corresponds to class switching between HepatoPredict classes associated with a very high or high predicted benefit of LT, respectively Class I and Class II [16]. Error B relates to a switch on HepatoPredict class with more impact in the LT benefit, i.e., a switch from a class with no benefit to a class with LT benefit (Class 0 to Class I or Class II) or from a class with LT benefit to a class with no benefit (Class I or Class II to Class 0). In general, error A was ∼1.6 times higher than error B (Figure 6E).

## Discussion

The HepatoPredict kit uses an algorithm that combines molecular data (gene expression levels of *DPT, CLU, CAPNS1,* and *SPRY2*) with clinical variables (tumor number, size of the largest nodule, and total tumor volume) to classify the patients in two different classes associated with the benefit of a liver transplant (Class I – very high confidence, and Class II – high confidence) or in Class 0 (no benefit of liver transplant predicted). Some products already exist in the market focused on the prognostic prediction of different tumors, such as breast [24] and prostate [25], but nothing specific for HCC is available. While the successful clinical validation of HepatoPredict kit using a retrospective cohort was previously published [16], the herein described new version of HepatoPredict algorithm presents several improvements. To start, the new HepatoPredict algorithm is associated with a higher PPV in Class I, which allows for the selection of very likely good prognosis candidates for LT, a potential advantage in geographies where the time in a LT waiting lists is very long. In addition, the new HepatoPredict algorithm presents a higher negative predictive value (NPV) when compared with different clinical criteria (such as Milan [4], UCSF [6], Up to seven [11], AFP [7], Metroticket 2.0 [5], TTV [13], and TTV AFP [12] criteria). The higher NPV of HepatoPredict translates into a higher probability of being correct in terms of detecting a bad prognosis patient when the kit result is Class 0 (no benefit of a LT). This reduces the misclassifications of patients that benefit from a LT and can avoid wasting a healthy organ in a patient that very likely will face HCC recurrence. This correct prognosis assignment was also corroborated by the results of multivariate analysis of the HepatoPredict algorithm, demonstrating a higher rate of type A errors (switch between Class I and II with benefit prediction) than type B errors (from good prognosis to bad prognosis and vice-versa). To further corroborate the clinical utility of the HepatoPredict kit additional retrospective studies are being planned enrolling patients’ cohorts from different geographic localizations and HCC etiologies and a prospective study (NCT0449983) is currently open and recruiting.

In the context of analytical validation of multi-target genomic assays (such as HepatoPredict kit), no evaluation guidelines covering all the relevant aspects required for the diagnostic setting are available. To fill this gap, different guidelines such as ISO201395-2019 [19], MIQE [20], CLSI-MM16 [22], CLSI- MM17 [21], and CLSI-EP17-A [23] were followed where applicable, to demonstrate that the HepatoPredict kit is a sensitive, specific, and robust test. Thus, the described analytical validation of the HepatoPredict kit is in accordance with standard assay validation processes. This type of approach has been previously used to validate similar prognostic [26–29] and diagnostic [30–32] tests and as a reference for analytical validation of the test in different molecular pathology laboratories.

In diagnostic settings, FFPE is the most commonly used technique for long-term conservation of clinical samples, since it preserves the proteins and vital structures within the tissue while it aids microscopic diagnostic examination, experimental research, and diagnostic/drug development [33]. FFPE samples were thus implemented for the HepatoPredict kit to simplify its adoption by molecular biology and pathology laboratories. The RNA extraction method from FFPE HCC samples was demonstrated to be repeatable (between duplicates) and reproducible (between operators). Regarding gDNA residual contamination, Cq values above 34 in RT-qPCR reactions targeting *Chr3* were observed in 1.15 and 9.19 % of the samples (for operator 1 and 2, respectively). However, the LoD for *Chr3* was determined at 33.95 Cq, meaning that above this Cq value, *Chr3* detection is likely invalid, suggesting that gDNA contamination during RNA extraction from HCC FFPE samples is very residual and approaching 0 % with HepatoPredict kit.

The RNA extracted from FFPE tissues is normally fragmented [34], thus, FFPE sections were digested with heat application (56 °C) and proteinase K to decrease RNA fragmentation and chemical modifications [35,36]. Moreover, the primers of the HepatoPredict kit were designed for the generation of short amplicons to increase gene detection rate [17,37,38], and gene specific reverse transcription and targeted cDNA amplification (1-step RT-qPCR) were performed to increase the accuracy and sensitivity of the RT-qPCR reactions [39]. Nevertheless, the HepatoPredict kit includes a sample quality control step comprising 1-step RT-qPCR reactions targeting *RPL13A* and *Chr3*. With this procedure, it is possible to determine if the extracted sample contains enough RNA (shown by the Cq value from *RPL13A*) and if gDNA contamination is present (reported by the Cq value from *Chr3*) [17] before proceeding with the kit protocol. This assessment is important because it assures the reproducibility and veracity of the experiments, avoids extra costs associated with the need of repeating the analysis and the wasting of precious tumor samples [20,40]. Considering that HepatoPredict analyses the expression level of 7 different genes, Cq values for each target, allowing all targets to be detected within their linear ranges and below their LoDs, were determined. Thus, an acceptable Cq range for the sample quality control was defined for the *RPL13A* gene: 18.32 to 28.26 Cq. Regarding *Chr3*, all Cq values above its LoD (33.95 Cq) are acceptable since they represent no gDNA contamination.

Regarding RT-qPCR reactions, primer specificity for each target included in the HepatoPredict kit was demonstrated. Although probes (TaqMan^Ò^ technology) are also included in the RT-qPCR reactions, they were not analyzed in the context of sequencing because they do not amplify PCR products and because their fluorescence is only released in the context of highly specific annealing to the target sequences in the PCR amplicons. Thus, if the Sanger sequencing proves that each primer pair-related PCR amplicon is specific and no unspecific PCR products are detected, each probe can only anneal to the amplified specific product. To further demonstrate that the RT-qPCR reactions included in the HepatoPredict kit could distinguish between target and non-target sequences, an inclusivity of 100 % was demonstrated for all targets, excluding the *DPT* gene which had an inclusivity of 95 %. This result was expected since the downregulation of the *DPT* gene in HCC has been demonstrated and can be already associated with HCC carcinogenesis and progression [41–43]. In contrast with diagnostic systems [31], the exclusivity of the RT-qPCR reactions was not studied since the HepatoPredict kit analyzes the expression level of genes that are not exclusively expressed on HCC cells. Nevertheless, to be analyzed by HepatoPredict kit, each HCC sample needs to be collected by expert clinicians (surgeons or radiologists) and subsequently analyzed at the microscopical level (H&E-stained tissue slides) by a certified pathologist, assuring the specificity of each HCC biopsy submitted to the HepatoPredict kit test. Furthermore, the sensitivity of the RT-qPCR reactions included in the HepatoPredict kit was determined by defining the LoD for each target, as well as the respective linear range. The amplification efficiency was also calculated for all targets being between 90 and 110 % as recommended by ISO 203095:2019.

The robustness of the qPCR reactions included in HepatoPredict kit was studied. It was demonstrated that qPCR reactions were robust, not being affected by small changes either in annealing temperatures and reagents’ concentration and volumes.

Moreover, precision studies, assessing both the repeatability and reproducibility of the qPCR reactions included in the HepatoPredict kit, demonstrated that the variability associated with normalized Cq values for *DPT, CLU, CAPNS1* and *SPRY2* genes was not dependent on a single factor (day, lot, or operator). Furthermore, the inter-assay SD within a sample is similar to the inter-assay SD between 18 samples, demonstrating the reproducibility of the assay independently of the sample used. The *DPT* gene was associated with a higher SD in all assays, nevertheless no differences were observed in *DPT* repeatability in comparison with the other targets. These results suggest that the lower reproducibility of the *DPT* gene (i.e., higher SD), may be associated with *DPT* lower inclusivity (95 %) due to *DPT* downregulation in HCC [41–43].

The robustness of the new HepatoPredict algorithm was also studied and the acceptable variation range for each variable was determined. It was demonstrated that *DPT* gene expression level and total tumor volume were the most sensitive variables. This was expected since in the XGBoost model (first level), the *DPT* gene expression level and total tumor volume had an information gain of 3.75 and 0.29 (respectively), while the other variables had an information gain of zero. Moreover, the SVM model (second level), also corroborated these results since the variables with higher SVM weights were *DPT* gene expression level, total tumor volume, and *CLU* gene expression level. Total tumor volume is related to the number of tumors and tumor diameter measurements; thus, errors in these variables will influence its value. A recent study described a mean error of 0.81 cm when measuring the tumor size using different magnetic resonance imaging pulse sequences [44]. This was reflected in an HepatoPredict type B error of 4.94 %, demonstrating that the prognostic test handles common measuring errors.

## Conclusions

Despite the introduction of perturbations mimicking real-life observed variations to the molecular and clinical variables, the prognostic information achieved with the HepatoPredict kit does not change. Prognosis variation is only expected under extreme and combined variations of multiple variables, unlikely to occur in real life. In addition, the technical validation procedures presented in this study can be used as a reference for the analytical validation of the HepatoPredict test in different molecular diagnostic laboratories. The performance of the presented analytical testing also demonstrates that the HepatoPredict kit can be easily integrated into routine molecular diagnostic procedures to accurately identify HCC patients more likely to benefit from a liver transplant, contributing to the implementation of a true precision medicine.

## List of Abbreviations

*AFP:*: alpha-fetoprotein
*DNA:*: Deoxyribonucleic acid
*CI:*: Confidence interval
*CLSI:*: Clinical laboratory standards institute
*Cq:*: Cycle threshold
*DCP:*: des-γ carboxyprothrombin
*FFPE:*: formalin-fixed paraffin-embedded
*gDNA:*: Genomic DNA
*HBV:*: Hepatitis B virus
*HCC:*: Hepatocellular carcinoma
*HCV:*: Hepatitis C virus
*ISO:*: International organization for standardization
*LoB:*: Limit of blank
*LoD:*: Limit of detection
*LT:*: Liver transplantation
*MIQE:*: Minimum information for publication of quantitative real-time PCR experiments
*NPV:*: Negative predictive value
*NTC:*: No template control
*N/A:*: Not applied
*p:*: p-value
*RNA:*: Ribonucleic acid
*RT-qPCR:*: real-time quantitative reverse transcription polymerase chain reaction
*SD:*: Standard deviation
*STD:*: Standard
*TTV:*: Total tumor volume
*UCSF:*: University of California San Francisco

## Declarations

### Ethics approval and consent to participate

The retrospective clinical study was approved by the ethics authorities (Comissão de Ética para a Saúde) from the Centro Hospitalar de Lisboa Central (Process number 144/2014). Being a retrospective study focused on tumoral tissue, the informed consent was waived by the same ethics authorities referred above. Samples were used in accordance with the Declaration of Helsinki.

### Consent for publication

Not applicable.

### Availability of data and materials

The datasets used and/or analyzed during the current study are available from the corresponding author on reasonable request.

### Authors’ contributions

MGR and JC conceived and designed the study. MGR and DP performed the experiments. MGR, LPF, JLN, and JC analyzed and interpreted the data. LPF drafted the manuscript. SS and HPM coordinated clinical sample collection. MGR, DP, JLN, JBPL, JC edited and revised the manuscript. All authors read and approved the final manuscript.

## Data Availability

All data produced in the present study are available upon reasonable request to the authors.

## Acknowledgements

The authors wish to thank to the patients, to *Neuralshift* and to the pathology team from the Curry Cabral Hospital, particularly Clara Rodrigues and António Figueiredo. Moreover, the authors particularly acknowledge the Biobank IRBLleida (PT20/00021) integrated in the Spanish National Biobanks Network and Xarxa de Bancs de Tumors de Catalunya sponsored by Pla Director d’Oncología Catalunya (XBTC), as well as the Biobank ISABIAL integrated in the Spanish National Biobanks Network and in the Valencia Biobanking Network for their collaboration.

