## Supplementary File 1 for "Analytical validation of HepatoPredict kit to assess hepatocellular carcinoma prognosis prior to a liver transplantation"

Supplementary File 1 – Demographic, clinical, and pathological characteristics of the cohort population

| | All patients (n = 162) |
|---|---|
| **Recipient characteristics** | |
| Male gender, n (%) | 148 (91.4) |
| Race (Caucasian), n (%) | 144 (95.4) |
| Age, years, median (IQR) | 55 (11.0) |
| MELD score, median (IQR) | 10.2 (4.7) |
| Waiting list, months, median (IQR) | 1 (2,5) |
| BMI, median (IQR) | 26.2 (5.8) |
| Ethanol intake, n (%) | 113 (72.4) |
| HBV infection, n (%) | 19 (14.0) |
| HCV infection, n (%) | 65 (47.8) |
| **Tumor related factors** | |
| AFP, median (IQR) | 7.4 (47.7) |
| Histological nº of tumors, median (IQR), range | 1 (1.0) 1-4 |
| Histological size of largest tumor, median (IQR) | 3.0 (2.0) |
| Microvascular invasion, n (%) | 24 (14.9) |
| Macrovascular invasion, n (%) | 16 (9.9) |
| Capsule, n (%) | 7 (5.1) |
| Poorly differentiated, n (%) | 22 (16.1) |
| Preoperative therapy, n (%) | 65 (47.4) |
| Incidental (%) | 0.0 |
| Within histological Milan Criteria, n (%) | 103 (64.0) |
| Histological total tumor volume ($cm^3$), median (IQR) | 14.1 (32.9) |
| Histological tumor volume ≤ 115 $cm^3$ | 151 (93.2) |
| **Operative data** | |
| Domino LT, n (%) | 77 (47.5) |
| Morbidity Clavien 3-4, n (%) | 33 (24.1) |
| Retransplantation, n (%) | 8 (5.0) |
| **Survival Data** | |
| Patients alive at 5 years, n (%) | 59 (36.4) |
| Recurrence at 5 years | 33 (20.4) |
| **Follow-Up** | |
| Follow-up, median (IQR), max. range | 63.5 (75.8) 195 |
