## Supplementary File 2 for "Analytical validation of HepatoPredict kit to assess hepatocellular carcinoma prognosis prior to a liver transplantation"

Supplementary File 2 – Inclusivity of the 8 targets included in the HepatoPredict kit.

| Targets | Number of detected samples / Total Samples | Inclusivity |
|---|---|---|
| *RPL13A* | 87/87 | 100 % |
| *GAPDH* | 19/19 | 100 % |
| *TBP* | 19/19 | 100 % |
| *DPT* | 18/19 | 95 % |
| *CLU* | 19/19 | 100 % |
| *CAPNS1* | 19/19 | 100 % |
| *SPRY2* | 19/19 | 100 % |
| *Chr3* | 11/11 | 100 % |

RT-qPCR reactions performed in duplicate by two different operators. Samples were considered detected when an acceptable Cq value (< 40) was obtained by the two operators.
