## Supplementary File 3 for "Analytical validation of HepatoPredict kit to assess hepatocellular carcinoma prognosis prior to a liver transplantation"

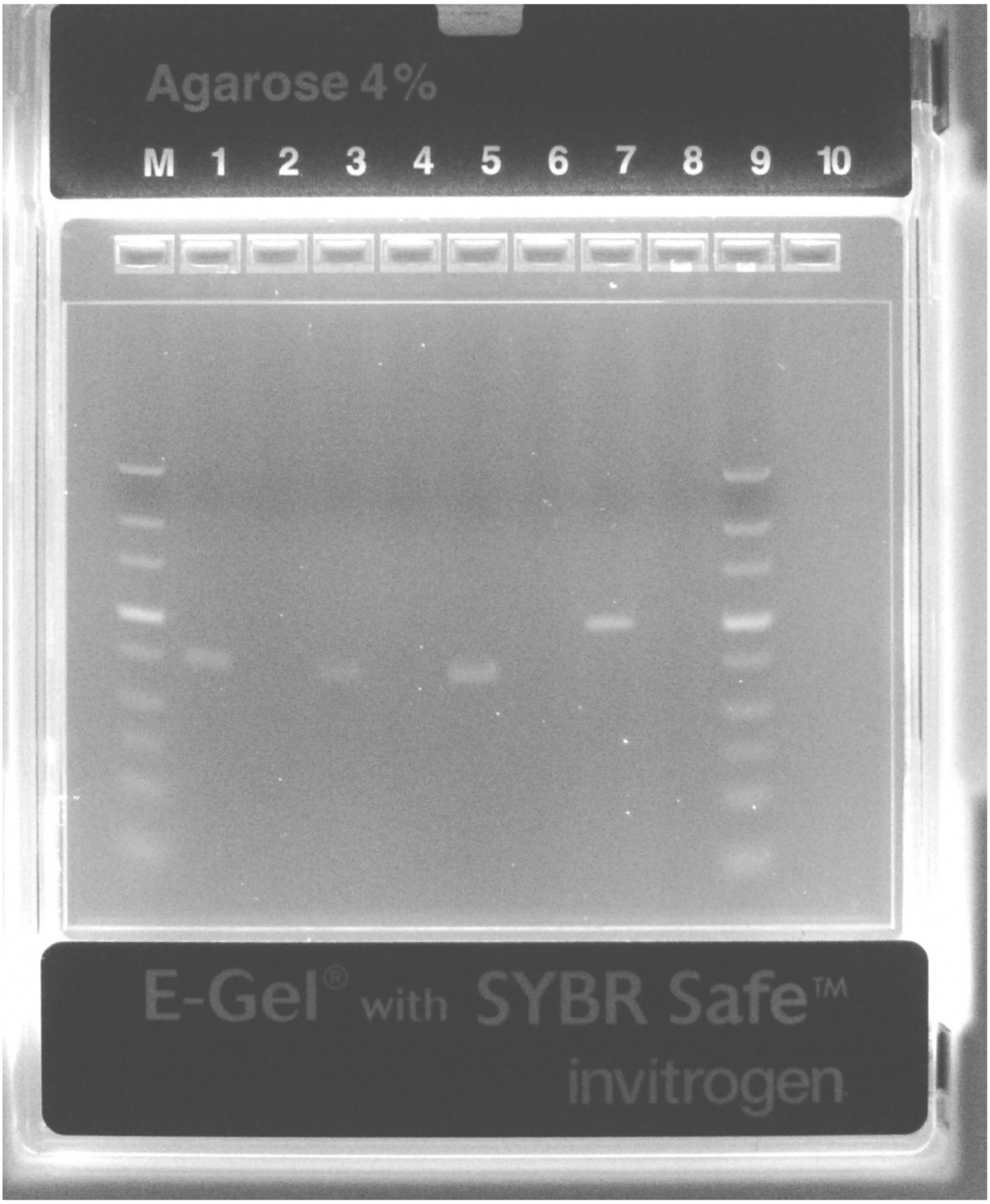


**Supplementary File 3 –** Uncropped agarose gel representing the DNA ladder (1st and 10th wells) and amplicons resulting from a RT-qPCR reaction using an RNA pool (2nd, 4th, 6th wells) or nuclease-free water (3rd, 5th, 7th wells) for *RPL13A* (2nd and 3rd wells), *TBP* (4th and 5th wells), and *GAPDH* (6th and 7th wells) genes. Additionally, an amplicon resulting from a qPCR reaction using gDNA or nuclease-free water for Chr3 is represented on the 8th and 9th wells (respectively). Finally, the 11th was loaded with nuclease-free water.
