## Supplementary File 4 for "Analytical validation of HepatoPredict kit to assess hepatocellular carcinoma prognosis prior to a liver transplantation"

Supplementary File 4 – Repeatability of the reference genes included in HepatoPredict kit

| Sample | Target | Repeatability (SD) |
|---|---|---|
| Sample A | *RPL13A* | 0.07 |
| | *GAPDH* | 0.08 |
| | *TBP* | 0.11 |
| Sample B | *RPL13A* | 0.07 |
| | *GAPDH* | 0.05 |
| | *TBP* | 0.06 |
| Sample C | *RPL13A* | 0.09 |
| | *GAPDH* | 0.06 |
| | *TBP* | 0.06 |

*SD, standard deviation.*
