## Supplementary File 5 for "Analytical validation of HepatoPredict kit to assess hepatocellular carcinoma prognosis prior to a liver transplantation"

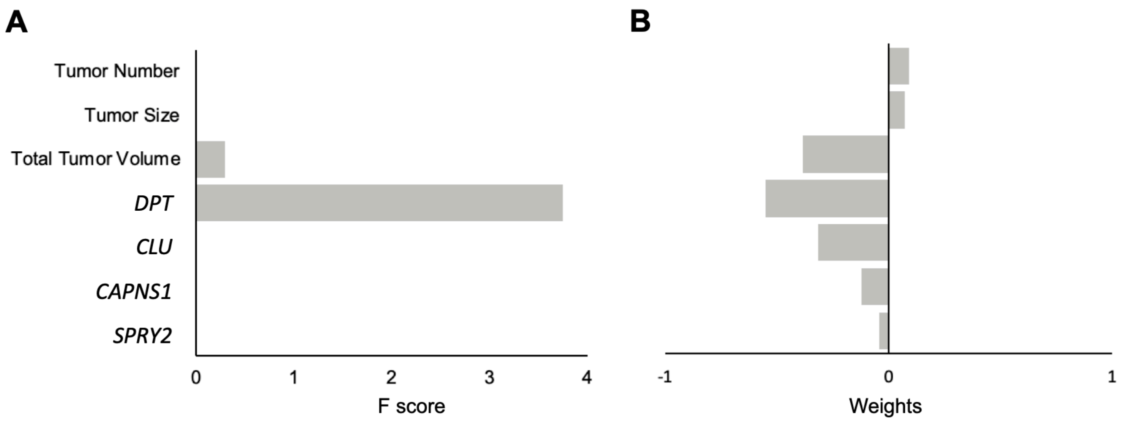


**Supplementary File 5 – Variables' weights in the new HepatoPredict algorithm.** Information gain of each variable for XGBoost model **(A).** Weight of each variable in the SVM model using a linear kernel function **(B).**
