## Supplementary File 6 for "Analytical validation of HepatoPredict kit to assess hepatocellular carcinoma prognosis prior to a liver transplantation"

Supplementary File 6 – Comparison of HepatoPredict predictive power with different clinical criteria for the identification of HCC patients suitable for liver transplantation. HepatoPredict used in highest sensitivity mode (Class I + Class II).

| | NPV | | Precision (PPV) | | Recall | | Accuracy | | FPR | | n |
|---|---|---|---|---|---|---|---|---|---|---|---|
| | Criteria | HP | Criteria | HP | Criteria | HP | Criteria | HP | Criteria | HP | |
| **Milan** | 36.21 % | 58.33 % | 84.47 % | 83.21 % | 70.16 % | 91.94 % | 67.08 % | 79.50 % | 43.24 % | 62.16 % | 161 |
| **USCF** | 40.68 % | 58.33 % | 87.25 % | 83.21 % | 71.77 % | 91.94 % | 70.19 % | 79.50 % | 35.14 % | 62.16 % | 161 |
| **Up to Seven** | 42.86 % | 56.00 % | 79.05 % | 83.21 % | 93.60 % | 91.20 % | 75.93 % | 79.01 % | 83.78 % | 62.16 % | 162 |
| **AFP** | 26.67 % | 41.67 % | 88.06 % | 90.00 % | 84.29 % | 90.00 % | 76.83 % | 82.93 % | 66.67 % | 58.33 % | 82 |
| **Metroticket 2.0** | 36.36 % | 45.45 % | 88.57 % | 90.00 % | 89.86 % | 91.30 % | 81.48 % | 83.95 % | 66.67 % | 58.33 % | 81 |
| **TTV** | 50.00 % | 52.63 % | 82.61 % | 85.83 % | 96.61 % | 92.37 % | 80.82 % | 81.51 % | 85.71 % | 64.29 % | 146 |
| **TTV AFP** | 37.50 % | 40.00 % | 90.14 % | 91.30 % | 92.75 % | 91.30 % | 84.81 % | 84.81 % | 100.0 % | 85.71 % | 79 |

HP (HepatoPredict new version), NPV (negative predictive power), PPV (positive predictive power), FPR (false positive rate).
